# Large Scale In-House Point-of-Care Manufacturing of Clinical Grade Autologous Anti-CD19 CAR T Cells in Saudi Arabia

**DOI:** 10.1101/2024.06.11.24308741

**Authors:** Walid Warda, Riad El Fakih, Nouf Alrasheed, Moumenah Alkhaldi, Noura Hamideddin, Hanan Alqudairy, Abdullah Alsuliman, Sheema Almozyan, Ayodele Alaiya, Alaa Osama Doubi, Haya S Alothaimeen, Ali D Alahmari, Feras A Alfraih, Mahmoud Aljurf, Hazzaa Alzahrani, Syed Osman Ahmed, Farhatullah Syed

## Abstract

Chimeric Antigen Receptor T (CAR T) cell therapy targeting the CD19 antigen represents a significant advancement in the treatment of various subtypes of CD19-expressing hematologic malignancies. Nevertheless, the current commercial CAR T manufacturing process is intricate, time-consuming, and costly, involving a supply chain that initiates at the treating center with mononuclear cell apheresis, followed by cryopreservation and shipping to and from a centralized third-party manufacturing facility. This process can introduce treatment delays due to logistical requirements. We hypothesized that point-of-care (POC) manufacturing of CAR T cells could reduce vein to vein time which is one of the major challenges with commercial CAR T processes. This study presents the outcomes of large-scale, clinical-grade, in-house POC CAR T manufacturing, including assessments of transduction efficiency, vector copy number, phenotype, and effector function of CAR T cells. Utilizing the CliniMACS Prodigy system, we processed mononuclear cells from fresh leukapheresis harvests of healthy donors. The cells underwent automated processing and activation with TransAct, a polymeric nanomatrix activation reagent containing CD3/CD28-specific antibodies. Following activation, the cells were transduced and expanded in the Centricult-unit under stabilized conditions, with automated feeding and media exchange. For transduction, we employed the Miltenyi Lentigen third-generation lentiviral vector, which delivers a second-generation CD19 CAR construct with a 4-1BB costimulatory domain and TNFRSF19 transmembrane domain. Our POC manufacturing data demonstrated the successful generation of large-scale CAR T cells, sufficient to prepare multiple doses within a shortened production period of 9 days, followed by cryopreservation for quality control assessment and batch certification for patient infusion. The transduction efficiencies, phenotype, and function of the Lentigen CD19 CAR T cells were comparable to previously published procedures, with overall T-cell yields sufficient for therapeutic application. CAR expression was evaluated by flow cytometry and vector copy number. In-process testing ensured the quality and suitability of CAR T cells in terms of sterility and cell number for the desired dose preparations. The final product was also subjected for the assessment of any replicative competent lentivirus. CAR-mediated target cell killing was assessed via co-culture assays, and cytokine release was measured by ELISA. To evaluate the shelf life of the cryopreserved product, stability and viability were checked at approximately 6 and 12 months. Our results indicate that in-house POC manufacturing at King Faisal Specialist Hospital & Research Centre (KFSH&RC) enables regulatory-compliant production of CAR T cells, eliminating the need for third-party shipping logistics.

## INTRODUCTION

Acute lymphoblastic leukemia (ALL) exhibits high survival rates in children (∼90%) (1) and 70% in relapsed cases (2), but adult survival rates are lower (50-60%) and worsen with age due to relapse and refractory disease. Novel agents like blinatumomab and inotuzumab show limited efficacy in relapsed B-ALL with <40% complete response rates and median survivals under 8 months (3, 4). In Saudi Arabia, adult ALL outcomes are around 60%, mainly due to relapse or refractory disease (5).

The advent of Chimeric Antigen Receptor (CAR) T cell therapy has revolutionized cancer treatment (6–8), providing new hope for patients with refractory or relapsed hematologic malignancies (6, 9, 10). However, its high cost, logistical complexities, and reliance on international manufacturing facilities hinder widespread adoption, particularly in Middle East and North African (MENA) region and GCC countries. This study proposes establishing the first point-of-care manufacturing facility for large-scale CAR T cell production in KFS&RC, Saudi Arabia, aiming to enhance accessibility, affordability, and efficiency of this therapy. The central hypothesis is that localized manufacturing can significantly reduce production costs, minimize logistical challenges, and expedite delivery of personalized cancer treatments, improving patient outcomes and fostering local biomedical expertise.

In preparation to establish In-house point-of-care (POC) manufacturing of CAR T cell for Phase I/II clinical trial (approval no. RAC#2191207), the project aim was to develop robust infrastructure for ATIMPs manufacturing (starting with CAR T cell production), for the first time in the kingdom, at KFSH&RC, creating a sustainable model for localized production replicable across the region. Key objectives were establishing a GMP-compliant facility equipped with advanced bioreactor and robust quality control systems, ensuring regulatory compliance in collaboration with the SFDA, and optimizing the production process to reduce costs. Additionally, the initiative focuses on training programs to build local expertise, developing an in-house advance CGT pipeline at R&D with a framework for clinical application within the KFSH&RC healthcare system, and integrating research to support continuous improvement and innovation in ATMP technologies. This effort will improve patient outcomes and position KFSH&RC as a leader in innovative cancer/gene replacement therapies and bio manufacturing, aligning with the goals of Vision 2030. To this end, once the lab was established, we hypothesized that in-house manufacturing of CAR T cells, on the automated Miltenyi CliniMACS Prodigy device, allows fast delivery of cells for the treatment of patients at KFSH&RC. Here, we describe for the first time in the kingdom a large-scale in-house clinical grade CAR T cell manufacturing results and transduction efficiency, sterility, impurities, vector copy number, phenotype, and effector function of CAR T cells. We utilized Miltenyi lentigen vector delivering a second-generation CD19 CAR construct with 4-1BB costimulatory domain and TNFRSF19 transmembrane domain and performed a large-scale production. Our point-of-care manufacturing data highlight the successful generation of large-scale CAR T cells at numbers sufficient to prepare multiple doses and an optional shortened duration of production time for 9 days followed by cryopreservation for QC assessment and batch certification for infusion into patients.

## METHODS AND MATERIAL

## 1. In-House POC Manufacturing of CAR T cells

ALL Lentigen CAR T cell manufactured at KFSH&RC Level 19 SIO7 (**Figure 1**) CAR T lab and the following steps includes (**Figure 2**):

**Figure 1.**
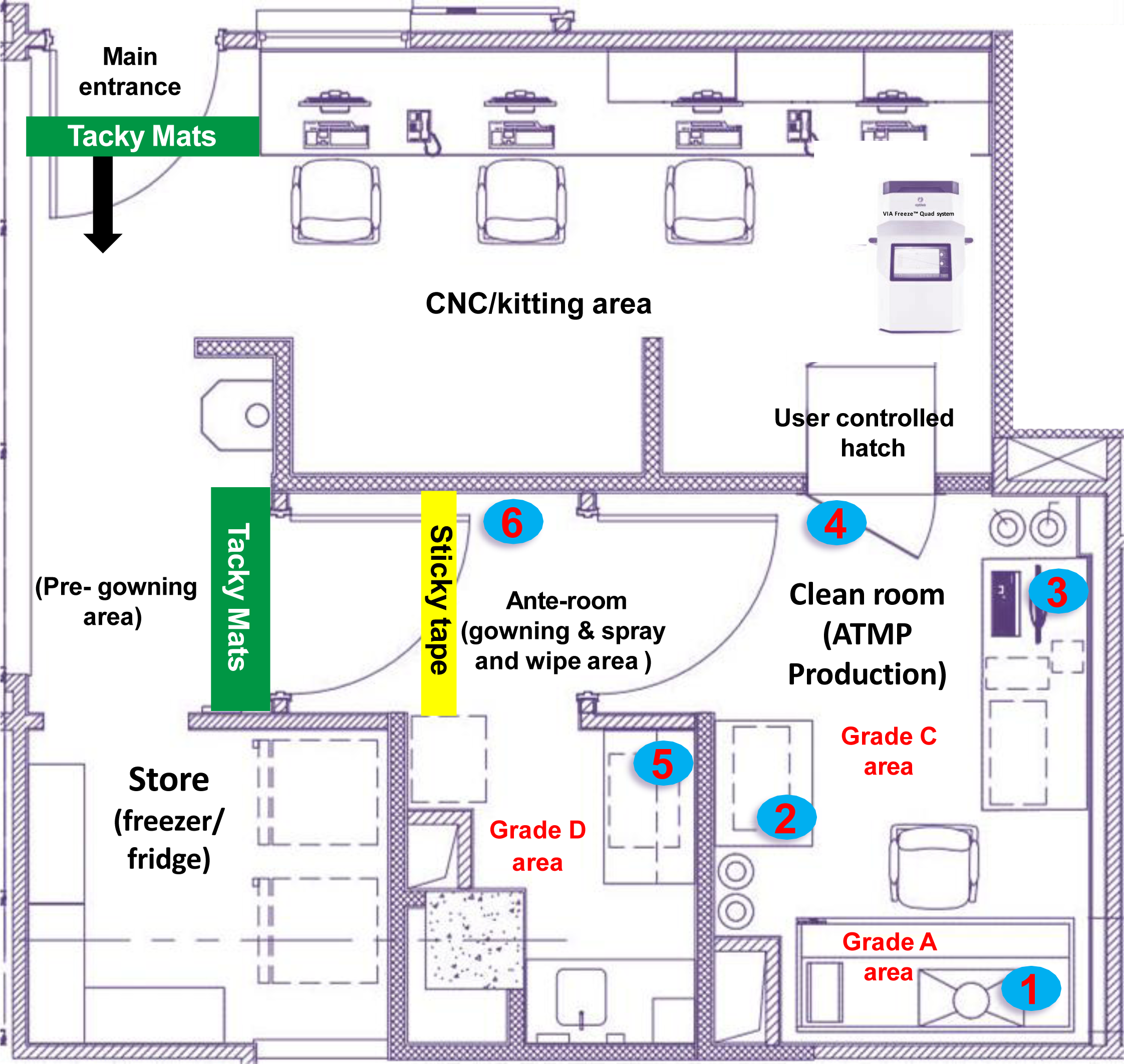
Schematic Diagram of GMP like ISO 7 Lab/class 10,000 Clean Room at KFSH&RC for POC ATIMP Production. The facility is designed to contain all the parameters such as; HEPA filters to ensure clean air circulation. Pressure differential to maintained in each area with alarms for deviations. Gowning Atrium Area for donning protective gear. Air Locks to prevents contamination between areas. Air Conditioners to maintain temperature and humidity and pressure monitoring systems. Tacky mats located near doors to trap contaminants. Sealed lighting to prevent dust accumulation. Protective gear requirements, Includes sterile gloves, bunny suit, booties, and hood etc. Stainless Steel bench for hygiene and ease of use. Cleanroom ISO7 filtration systems with Class 10,000 filtration provides 15-25% filter coverage with a minimum of 60-65 air changes per hour. The schematic illustrates the layout of the Controlled Not Classified (CNC) kitting area, the gowning area, and the production area, which includes six designated locations for environmental monitoring (EM) using Tryptic Soy Agar (TSA) plates. These locations numbered 1 to 6 in the schematic, with location 1 representing the cleanest area and location 6 representing the least clean area. During the manufacturing process, TSA plates are exposed in these areas for up to 4 hours to monitor environmental conditions. This procedure ensures the maintenance of aseptic facilities and the proper operation of aseptic production equipment in clean room environments. EM monitoring is a mandatory test as per current Good Manufacturing Practice (cGMP) regulations. **Note:** Microbiological cleanliness levels “In Operation” (Settle Plate diameter 90 mm; CFU/4 hrs): Location 6. Controlled support area (cfu/plate: <100); Location 5. Controlled processing area 1 (cfu/plate: <50) Location 4. Controlled Processing area 2 (cfu/plate: <50); Location 3. Controlled Processing area 3 (cfu/plate: <50); Location 2. Aseptic Processing area (cfu/plate: <5); Location 1. Aseptic Core area (cfu/plate: <1).

**Figure 2.**
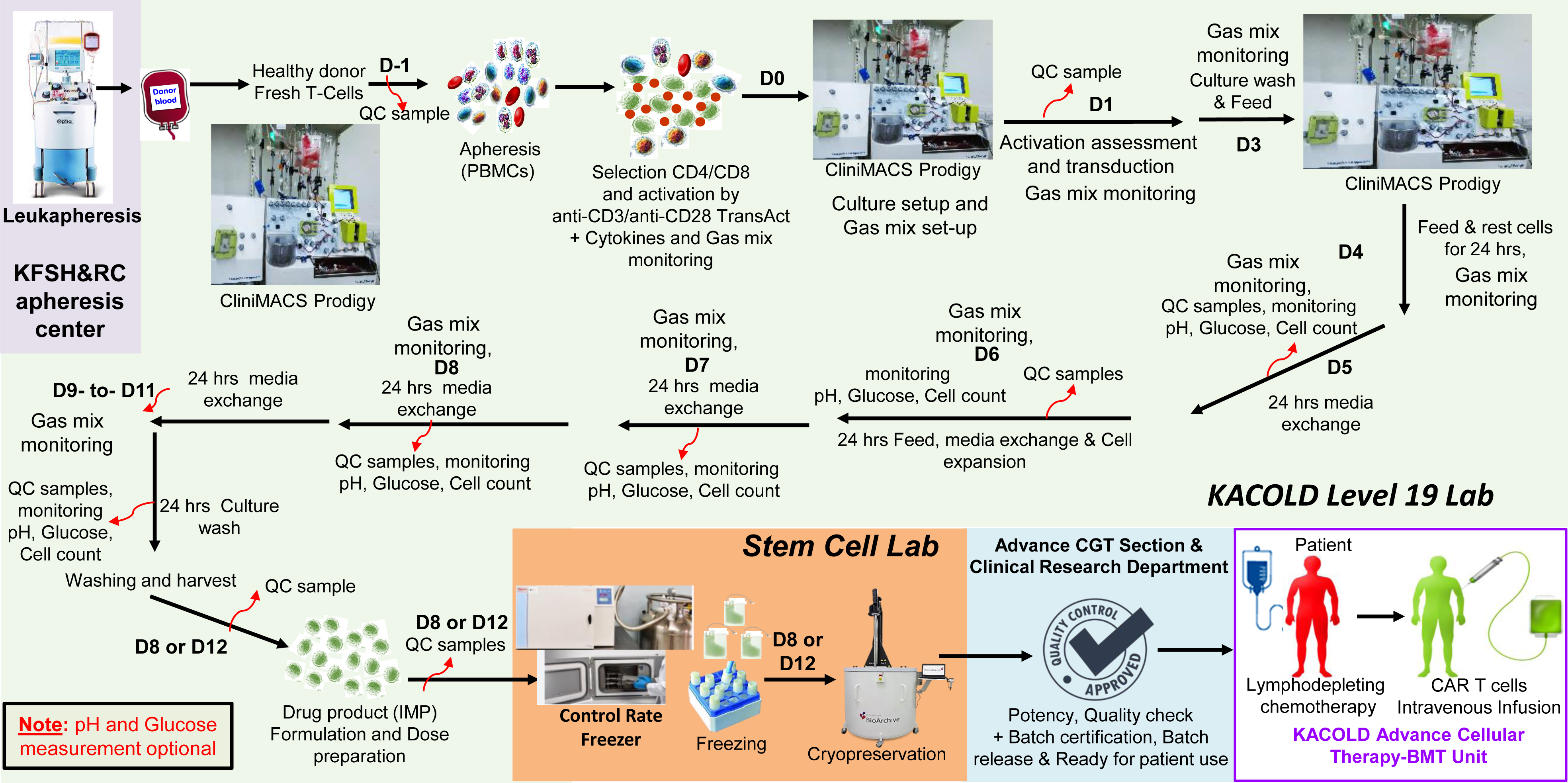
In-house POC Manufacturing of CAR T cell Process Map. Schematic shows the manufacturing steps for ALL Lentigen CAR T cell and Highlights the multidisciplinary collaboration for In-house point-of-care production of ALL Lentigen CAR T cells using CliniMACS Prodigy.

### 1.1 Step 1: Donor Leukapheresis (day −1)

This project was approved by the ethics committee of the KFSH&RC, Riyadh (approval no. RAC#2191207). The participant gave written informed consent. Peripheral blood mononuclear cells were isolated after leukapheresis acquired from healthy volunteer donors at blood bank, KFSH&RC.

Within 30 days prior to the leukapheresis, the clinical trial donor was tested for HBsAg, anti-HBcore, Hepatitis B DNA, Anti-HCV-Ab, Anti-HTLV 1+2 and Syphilis serology at KFSH&RC, under hospital regulations. PBMCs were harvested from the donor by unstimulated (no G-CSF used) apheresis (referred to as leukapheresis) at KFSH&RC apheresis lab. The collected cells were initially taken to the KACOLD level 19 CAR T lab for generation of the ATIMP, or delivered directly to KFSH&RC in the case of unrelated donor cells. If cryopreservation of leukapheresis is necessary this will be performed by DPLM labs at KFSHRC as a routine procedure. A sample of the apheresis is analysed using a Sysmex (CBC) automated haematology analyser. This determines the leucocyte and lymphocyte count as well as red cell contamination as shown in the **figure 3B**.

**Figure 3.**
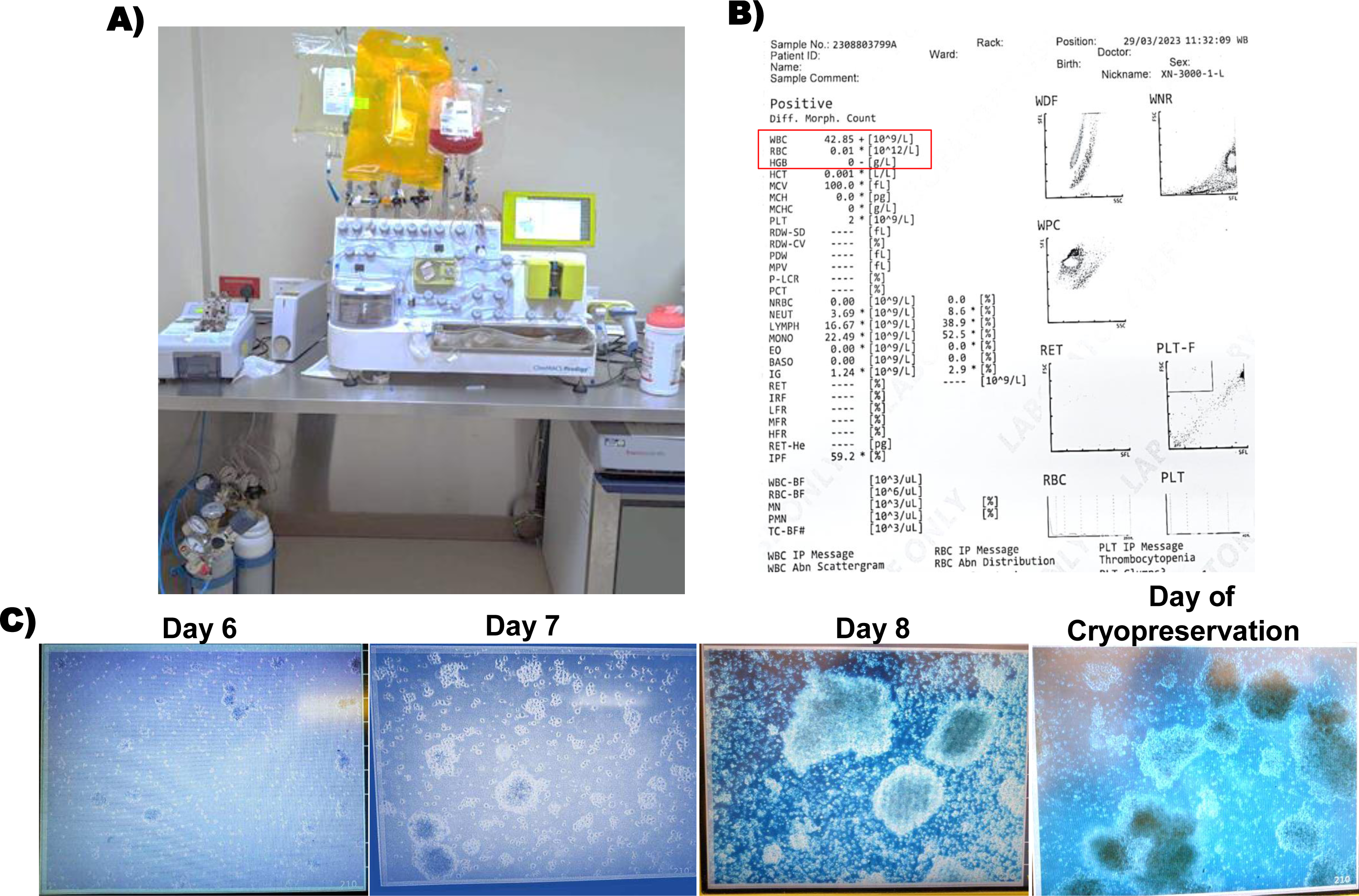
Large-scale Clinical Grade CAR T cell Manufacturing on CliniMACS Prodigy. **A)** The Single-use Tubing Set TS520 enables sterile handling of cell products within a single container. Integral to the tubing set is the CentricultUnit, which allows for stable temperature and gas mixture maintenance, as well as the capability to wash cells during the procedure without removing them from the device. **B)** Sysmex analysis results of the starting material display a complete blood count. **C)** The automated TCT process promotes efficient expansion of CAR T cells, as evidenced by micrographs captured by the internal camera of the CliniMACS Prodigy at various time points indicated in the figure.

### 1.2 Step 2. Start of Culture, Activation of PBMCs (day 0)

Approximately 0.7-1×10^8^ WBC cells are needed to start the T cell transduction (TCT) in the CliniMACS Prodigy. On day 0 cell, count is determined by FACS/ or on a Sysmex automated cell counter or other validated counting methods and a sample was taken for FACS analysis and sterility tests (BacT/Alert or BACTEC). The cell bag containing the fresh product cells, the media (TexMACS, 3%human AB-Serum and 20ng/mL hIL-2) and the activation reagent TransAct are connected to the CliniMACS Prodigy. TransAct is used in a dilution of 1:200 for MACS GMP TransAct CD3 Reagent and 1:400 for MACS GMP TransAct CD28 Reagent. The cultivation is defined by programing the activity matrix, which includes timing of transduction, culture wash and media exchange. Adaptation of media replacement and duration of expansion can be performed during the process and is dependent on cell expansion in the culture.

### 1.3 Step 3 Transduction (day 1)

Activated PBMCs are transduced on D1 of the manufacture process, the cells are transduced with Lentigen aCD19 - CAR LTG1563 lentiviral vector developed and provided by Lentigen, a Miltenyi Biotec company. All experiments described in this manuscript used a scFv FMC63-based targeting domain, CD8-derived hinge region, TNFRSF19-derived transmembrane region, 4-1BB/CD137 costimulatory domain, and CD3-zeta chain intracellular signaling domain, which was developed and provided by Lentigen, a Miltenyi Biotec company (Gaithersburg, MD, United States).

At the programed time for transduction, the operator is instructed to connect the vector bag containing the viral vector to the TS520 tubing set. The 10mL of diluted viral vector is then taken into the culture chamber and the vector bag is washed with a further 20mL of complete media which is also transferred to the culture chamber.

### 1.4 Step 5: Culture Wash and Cell Culture Expansion (day 3-to-day 7/11)

In the activity matrix, an automated culture wash is programed for between 48 hours post activation of the cells. The automated culture wash involves a complete change of the cell culture media and removes lentiviral vector and excess activation reagent. A sample was taken from the culture chamber using the automated sample function on D4 to D7. This sample is used to assess the cell count and other parameters (pH and glucose, optional) depending on the cell count shaker 2 (>3×10^6^/mL) or shaker 3 (>4×10^6^/mL) is activated. A specification of ≥ 15% transduction efficiency on day 6 is set for the manufacture to proceed. If this specification is not met, then the process is terminated.

### 1.5 Step 6: Harvest and Cryopreservation QC Sampling (day 8/12)

On day 8 or day 12 depending on the required cell number, product is harvested from the CliniMACS Prodigy following the end of culture activity using formulation buffer, CliniMACS 1% HAS or equivalent ready to use cryopreservation media. Samples from the harvested cells were taken for CD45/CD3+ and CAR+ transduction efficiency and CD45+/CD3+ purity. The cells were cryopreserved in doses based on the CD45+ CD3+ CAR+ T cells. The cells were cryopreserved in cryosure Bags using control rate freezer (CRF). Once the CRF is successful, the cryopreserved doses were transferred to vapor phase liquid nitrogen for long-term storage (**Figure 2**).

## 2. Analytic Procedures

All assays are validated, and where possible, performed in CAP accredited KFSH&RC laboratories according to the established standard SOPs. All assays are appropriately quality controlled.

### 2.1 Flow cytometry for Cell Counting, Viability and Transduction

Briefly, cell product is washed and incubated with antibodies that recognise CD3, CD4, CD8, CD45, CD19, CD16/56, CAR19 and 7-AAD etc. in flow tubes. Non-transduced cells are washed and stained in parallel. Viability is calculated using 7-AAD in tubes with CD45 and CD3.

### 2.1 Sysmex Count

The Automated Hematology System (Sysmex) uses fluorescent flow cytometry and hydrodynamic focusing techniques to differentiate cell types in whole blood and cell suspensions. It is used routinely for the processing of patient samples at KFSHRC.

### 2.2 Sterility by Gram Stain

The test sample is smeared onto a slide and stained with Gram stain. The peptidoglycan cell membrane on Gram positive bacteria, if present, will stain blue, Gram negative bacteria appear red. This assay is used routinely by KFSH&RC-microbiology laboratory to test supernatants for the presence of bacteria in other apheresis products.

### 2.3 BacT/Alert or Bactec Blood Culture System for Sterility Testing

The BacT/Alert or BACTEC sterility assay are standard bacteriological techniques carried out routinely by KFSHRC microbiology laboratory. Anaerobic and aerobic BacT/ALERT blood culture bottles were inoculated with the test sample in the clean room facility and are incubated for 7 days. Bacterial growth is determined using the BacT/ALERT or validated equivalent. This system is used routinely by KFSH&RC Microbiology to test other apheresis products for the presence of gram-positive bacteria in other cell therapy product prior to patient infusion.

### 2.4 Mycoplasma Testing

Mycoplasma infection is excluded by Mycoplasma genus test, which is a standard PCR or real time PCR based method. The test article is cells from the culture process. The PCR method used employs primers targeting for the presence of Mycoplasma genus (including but not limited to: bovis, fermentans, genitalium, hominis and penetrans) DNA. The test is validated by our KFSH& RC team at Microbiology and KFSH&RC R&D laboratories.

### 2.5 Endotoxin Testing

The test method is the Kinetic Turbidimetric method and is as described in the European Pharmacopoeia Monograph. Cell supernatant from process culture is mixed with the reconstituted LAL reagent, placed in the incubating microplate reader, and automatically monitored over time for the appearance of turbidity. The time required before the appearance of turbidity (Reaction Time) is inversely proportional to the amount of endotoxin present. That is, in the presence of a large amount of endotoxin the reaction occurs rapidly; in the presence of a smaller amount of endotoxin the reaction time is increased. The concentration of endotoxin in unknown samples can be calculated from a standard curve. Dilutions of cell samples are performed to determine an optimum dilution within the release limit of <2.0EU/ml for ATIMP products. This test is well established and routinely done at KFSH&RC toxicology laboratory for testing other apheresis products.

### 2.6 Cell Lines and Master Cell Bank

Raji and Daudi (B-cell malignant cell lines) and K562 cell line were originally purchased from the American Type Culture Collection (CCL-86, CCL-213 and CRL-3344 respectively). The cell lines were maintained in R10 medium (RPMI medium 1640 supplemented with 10% FBS, 1% GlutaMAX supplement (Gibco; Cat# 35050–079), 1% Pen/Strep (Gibco; 15140–122) at 37 °C in 5% ambient CO_2_. All cell lines were tested for mycoplasma contamination every batch production (Lonza; Cat# LT07-710). To generate a master cell bank, we cultured each cell line under optimal conditions, performed multiple passages to ensure stability, and verified cell line identity (CD19 expression in flow cytometry) and then cryopreserved aliquots at low passage numbers in liquid nitrogen for long-term storage and future use in potency assays.

### 2.7 Flow Cytometry for Drug Product

The flow cytometric analysis procedure for various parameters has been internally validated by the manufacturing team using the MACSQuant® Analyzer 10 Flow Cytometer and BD LSRFortessa™ High-Parameter Flow Cytometer. Validation was performed on both instruments by two independent operators where specified. Peripheral blood cells from leukapheresis were phenotyped using CD45 VioBlue, CD3 FITC, CD4 VioGreen, CD8 APC-Vio770, CD14 APC, CD19 PE-Vio770, CD56 PE, CD16 PE, and 7AAD (Miltenyi Biotec) to establish the following populations: CD45+ lymphocytes, CD4+ T cells, CD8+ T cells, CD4+CD8+ T cells, CD3+ T cells, CD14+ monocytes, CD19+ B cells, CD56+CD16+ NK cells, and CD3+CD56+CD16+ NKT cells. CAR T cells were phenotyped with CD19 Fc Chimera protein for 10 minutes at 4°C. Cells were then washed and stained with an anti-biotin-PE secondary antibody for 10 minutes at 4°C, along with CD45 VioBlue, CD4 VioGreen, CD8 APC-Vio770, CD3 FITC, and 7AAD (Miltenyi Biotec). Un-transduced T cells were used as a negative control. Transduction efficiency was identified as the percentage of CAR+ among 7-ADD-CD45+CD3+ cells. Samples were analyzed using the LSR Fortessa (BD Biosciences) and and MACSQuant® Analyzer 10 Flow Cytometer. Data data were processed with FACSDiva 8.0 software (BD Biosciences)), and MACSQuant software (Miltenyi Biotec).

### 2.8 Vector Copy Number

The vector copy number (VCN) assay is intended to determine the average copies of integrated lentiviral vectors per cell genome after ex vivo transduction of human cells. After isolating genomic DNA from the transduced cells using DNAeasy Blood and Tissue kit (Qiagen, 69506), the copies of the lentiviral gag gene are determined in relation to the human reference gene PTBP2 by quantitative PCR. The MACS COPYcheck Kit (Miltenyi Biotec, 130-128-157) provides specific primers for amplifying the lentival gag gene and the human reference gene PTBP2 using real time quantitative PCR.

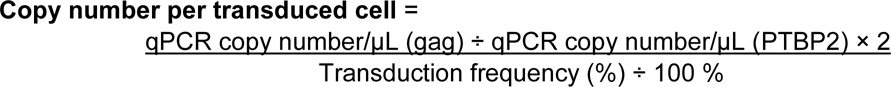

### 2.9 Replication Competent Lentivirus

The MACS LENTIcheck Kit (Miltenyi Biotec, 130-128-760) was used to amplify an amplicon of the lentiviral envelope gene sequence VSVg in real time quantitative PCR. The determined value to the power of 10 gives the copy number of VSV-G in the respective sample. The linear equation of the standard is: Ct = slope × log(copies/µL) + Y-intercept. The mean Ct value for each triplicate sample is used to determine the log(10) of the number of copies according to the respective standard curve. Results < 40 Ct are positive.

### 2.10 Cytotoxicity Assay

Cytotoxicity assays were performed by co-culturing CAR T cells with target cells at various effector to target ratios. Briefly, 100,000 target cells (K562, Raji or Daudi) were plated per well of a 96-well plate and various ratios of CAR T cells were added. At the end of the culture, cells were stained with DAPI for viability, CD3 for the identification of the effector cells. After 15 min, Flow cytometry Absolute count standard (Thermo Fisher) were added to the tube and cells were analyzed by flow cytometry. Absolute numbers of live target cells at the end of the assay were calculated by comparing the ratio of bead events to cell events. The percentage of cytotoxic activity was determined by dividing the absolute number of target cells at the end of the culture with the absolute number of target cells at the beginning of the experiment.

### 2.11 Interferon-γ (IFN-γ) Intracellular Staining

Effector cells (un-transduced T cells and CAR T cells) were co-cultured with either K562, Raji or Daudi cells at a E:T ratio of 1:4 for 6 hours in presence of Golgi Plug (BD Bioscience). Effector cells alone were used as negative control. At the end of co-culture, cells were stained with anti-CD3, CAR detection recombinant protein and 7-AAD. After washing, cells were fixed/permeabilized using BD Cytofix/Cytoperm Kit (554714) and cells were stained with anti-IFN-γ PE and anti-TNF-a PE-Cy7. Data were acquired on an LSRII flow cytometer (BD Biosciences). BD FACSDiva software (BD Biosciences) was used for further data analysis.

### 2.12 IFN-γ Secretion Assay

Effector and target cells were co-cultured with either K562, Raji or Daudi cells at a ratio of 1:4 in TexMACS medium for 24 hours. Co-culture supernatants were collected and stored at −80 °C. The concentration of IFN-γ in co-culture supernatants were determined by ELISA using Quantikine® Human IFN-γ Immunoassay kit (R&D systems, DIF50C). Briefly, plates were coated with a mouse anti-human antibody specific for each protein of interest. Samples of unknown concentration were added to the plate. Biotinylated mouse anti-human IFN-γ antibody was added, followed by streptavidin–horseradish peroxidase. Finally, a 1:1 solution of hydrogen peroxide and 3,3′,5,5′-tetramethylbenzidine was added to plates to induce a colour change, and the reaction was stopped by addition of 2 m sulphuric acid. Between each step, plates were washed three times in wash buffer. The final absorbance of each sample was read at 450 nm and optical densities were measured.

### 2.13 Fluorescent Labeling of Cells for Trogocytosis Detection

Target cells were labeled with the lipophilic membrane stain DiI (1,1′-dioctadecyl-3,3,3’,3’-tetramethylindocarbocyanine perchlorate; λEx/λEm = 549/565 nm) and effector cells were labeled with another lipophilic membrane stain DiD (1,1′-dioctadecyl-3,3,3′,3′-tetramethylindodicarbocyanine, 4 chlorobenzenesulfonate salt; λEx/λEm = 644/665 nm) according to the manufacturer’s instructions (Vybrant Multicolor Cell-Labeling Kit, Molecular probesm, V-22889). Briefly, cells were resuspended in serum-free RPMI-1640 medium to a concentration of 1 × 10^6^ cells/mL. Five μL of the dye solution (1 mM) was added to each 1 mL of cell suspension and incubated for 20 min at 37 °C in the dark and then centrifuged at 300× *g* for 5 min at RT. The stained cell pellet obtained was further subjected to three rounds of centrifugation in growth media to remove unbound dye. Cells were co-cultured according to the protocol described above for IFN-γ release assay.

### 2.14 Statistical Analysis

Unpaired two-tailed t-tests were used for comparison of two groups. One-way ANOVA was used when more than two groups were analyzed for statistical significance. All statistical analyses were done using GraphPad Prism 8.3.0 (GraphPad Software, San Diego, CA, United States). Statistical significance was given as ∗, ∗∗, ∗∗∗ by p-values less than <0.05, <0.001 and <0.0001, respectively.

## 3. Quantitative Expression Proteomics by Label-Free Liquid Chromatography/Mass Spectrometry

### 3.1 Sample Preparation for Proteomics

The whole cell lysate of complex protein mixture derived from Un-transduced T cells & CD19 CAR T cells sample groups was processed for in-solution trypsin-enzyme digestion as previously described with minor modifications (11). Briefly, cell pellets derived from samples were solubilized in RapiGest SF buffer (Waters, Manchester, UK) and protein concentration determined for each sample. Equivalent of 100μg of total protein from each sample, was denatured at 80 ◦C for 15 min. The samples were then reduced in 10 mM DTT at 60^◦^C for 30 min and finally alkylated using 50 mM Iodoacetamide, for 40 mins in the dark at room temperature. All samples were digested using sequence grade trypsin (Promega) at a ratio of 1:50 (w/w) and left overnight at 37^◦^C.

### 3.2. Label-Free Quantitative Expression Proteomics

Global expression Protein identification was done using one-dimensional (1D) - Nano Acquity label-free liquid chromatography coupled with Synapt G2 HD Mass Spectrometry instrument (Waters, Manchester, UK). The pre-analysis instrument tune parameters were done using the MassLynx, as previously described (11, 12). All samples were run in triplicates on Trizaic Nano Source, in resolution and positive ion mode nanoESI in the AcquityTM HSS T3 85 um × 100 mm column (Waters, Manchester, UK). The MS data were acquired with a gradient acquisition run time of 120 min in an m/z range of 50-2000 Da using data-independent acquisition (DIA)/ion-mobility separation experiments (HDMSE), (MassLynx version. 4.1, SCN833; Waters, Manchester, UK).

### 3.3. Data Analysis

The acquired raw MS data were processed for protein identification, by database searching in non-redundant UniProt/SwissProt specie-specific (Homo sapiens) using Progenesis QIfP V4.0, program (Waters, Manchester/Nonlinear, Newcastle, UK) as previously described [AlZahrani 2022, Alkhayal 2023]. Only proteins with marked expression changes with a p-value < 0.05 and at least ≥ 2-fold were considered statistically significant.

## RESULTS

## 1. Establishing an ISO8/ISO7 GMP Laboratory for ATIMP manufacturing at KFSH&RC, Saudi Arabia

Setting up an ISO8/ISO7 GMP-compliant laboratory for in-house POC manufacturing involves meticulous planning and adherence to regulatory standards. The following outlines the key steps and considerations for this project:

### 1.1 Facility Design

The first step was establishing a GMP facility that meets ISO8/ISO7 standards (**Figure 1**). This included:

- Cleanroom Environment: The laboratory is equipped with controlled environments to minimize contamination. As ISO8 and ISO7, classifications require stringent control of airborne particulate levels. For preventing cross-contamination coming from the adjacent ante-room, a positive clean room-pressure of about 15 Pascals (Pa) is essential for airflow from higher cleanliness to a lower cleanliness graded area. The ante-room was designed with a positive pressure of 2.5 Pa to eliminate contamination from the pre-gowning area which is classified as ISO8 room. Moreover, each room is equipped with a room pressure monitor which alarms when the pressure is interrupted.
- HVAC Systems: Robust heating, ventilation, and air-conditioning (HVAC) systems to maintain air quality, temperature, and humidity. The air change per hour (ACH) in the clean room and ante-room is 65 and 30, respectively. Continuous monitoring systems are also in place for recording the data from each area.
- Laboratory Layout: Designed the layout that promotes a workflow to prevent cross-contamination, such as separate areas for raw material handling, cell processing, and product storage.
- Utilities and Equipment: Ensuring reliable utilities (CO_2_, and medial gas) and installed calibrated equipment such as biosafety cabinet, centrifuge, incubator (optional) balance and automated cell processing systems etc. All equipment installed in the laboratory undergoes Planned Preventive Maintenance (PPM) periodically. The laboratory is readied with a continuous power supply, eliminating interruption to critical laboratory operations.

### 1.2 Regulatory Compliance and GMP Standards

Compliance with local and international regulatory standards is critical. This includes:

- Regulatory Approvals: Obtaining necessary approvals from SFDA and aligning with international standards such as those set by the FDA and MHRA/EMA.
- GMP Guidelines: Adhering to GMP guidelines for facilities, equipment, personnel, and processes to ensure product safety, efficacy, and quality.
- Documentation: Establishing thorough documentation practices, including validation and qualification protocols for equipment and processes (**Figure 2 and Table 1**). Also, utilizing robust tracking forms and labeling system which is critical to maintain the chain of custody and identity of the ATIMP product.

**Table 1.**
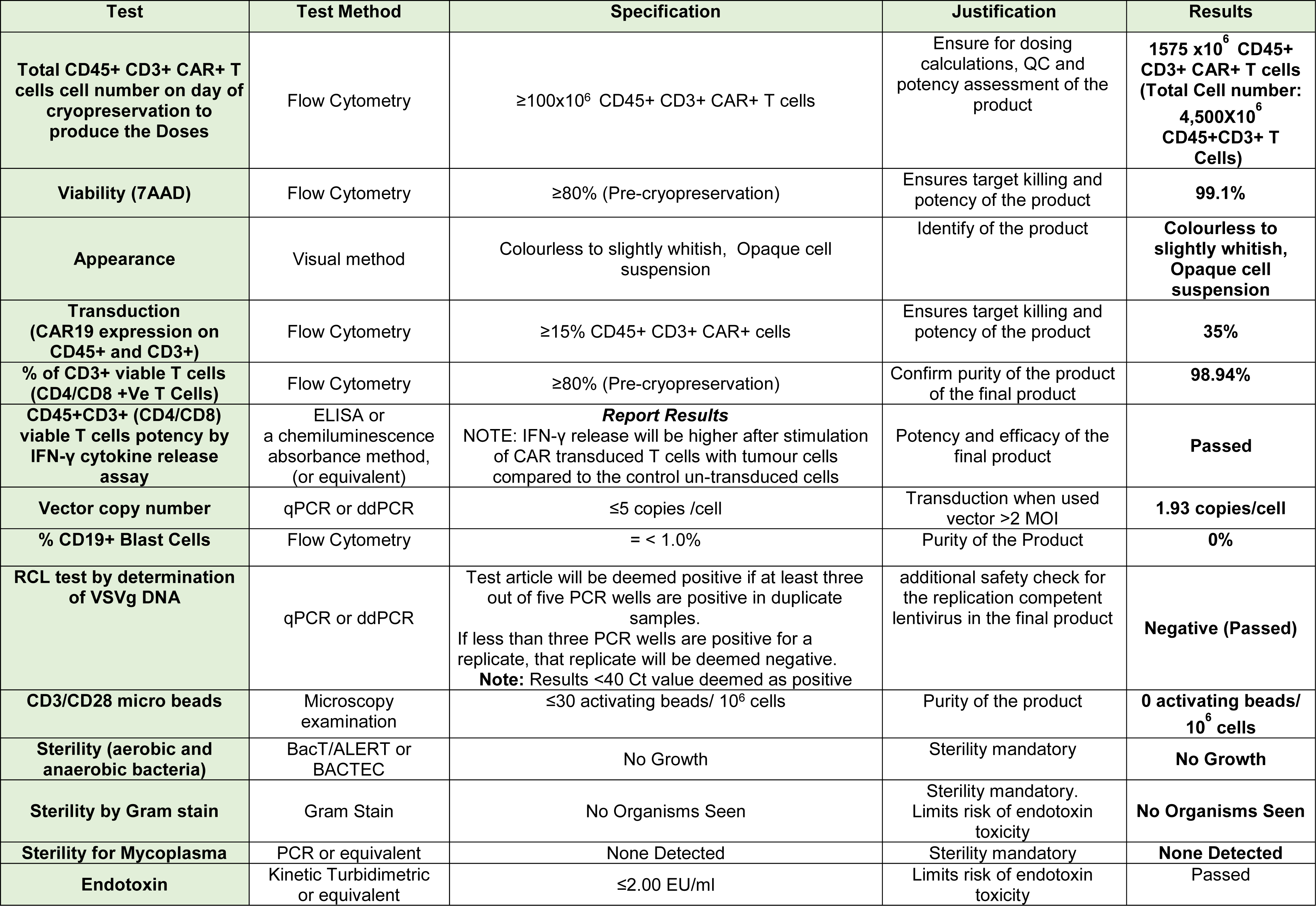
Specifications for drug product release criteria (ALL LentigenCAR T cells).

### 1.3 Training and Competency Development

Staff training is a legal requirement for ATMP production, therefore below mandatory training was implanted during this manufacturing procedure:

- GMP Training: Training programs on GMP principles, procedures, and regulations.
- Technical Skills: Hands-on training for staff on CAR T cell manufacturing techniques, including cell culture, transduction, expansion, and cryopreservation.
- Quality Control: Training in quality control procedures to ensure the integrity and potency of CAR T cell products.

### 1.4 Establishing a Quality Management System

A robust QMS is essential for maintaining high standards of quality. This involved (**Table 1 and 2**):

- Quality Assurance (QA): Developed QA processes to monitor and evaluate all aspects of the manufacturing process.
- Quality Control (QC): Implemented QC tests to verify product quality at various stages of production.
- Standard Operating Procedures (SOPs): Detailed SOPs were established for all procedures and processes to ensure consistency and compliance.
- Continuous Improvement: Established mechanisms for continuous improvement, including regular cleaning, environmental monitoring, audits, feedback loops, and corrective action plans.

### 1.5 Operationalizing the ISO7 Laboratory

Once the facility was ready and the staff was trained, the lab needs to be operationalized. Steps included:

- Process Validation: Conducted process validation to demonstrate that the manufacturing processes produce product meeting predetermined specifications (**Table 1 and 2**).
- Validation Runs: Performed initial production run to troubleshoot and refine processes (**Table 1 and 2**).
- Inventory Management: Setting up systems for raw material and reagent inventory management to ensure uninterrupted production.
- Scheduling and Workflow Management: Developed efficient scheduling and workflow management systems to optimize lab operations and resource utilization.

### 1.6 Establishing In-house Analytical Tests

In-house analytical tests are crucial for ensuring the quality and safety of CAR T cell products (**Figure 2 and Table 1**). These included:

- Assay Development: Developed and validated assays for identity, purity, potency, and safety testing of CAR T cell products.
- Flow Cytometry: Established phenotypic characterization and functional assessment of CAR T cells.
- Molecular Testing: Implemented molecular assays such as VCN, RCL and sequencing for transgene integration and expression analysis.
- Microbial Testing: Implemented all the required microbial testing to ensure the sterility of cell products (**Table 1 and 2**).

### 1.7 Writing SOPs and Documentation

SOPs are critical for CAR T manufacturing for ensuring consistency and compliance for analytical methods. Key aspects included:

- Process SOPs: established SOPs for each step of the CAR T cell manufacturing process, from raw material handling to final product cryopreservation.
- Analytical SOPs: Developed SOPs for all analytical tests performed in-house.
- QA/QC SOPs: Created SOPs for quality assurance and control processes, including documentation, auditing, and reporting procedures.
- Safety SOPs: Established SOPs for safety protocols, and handling viral vector to protect staff and ensure a safe working environment.

## 2. Automated Manufacturing of Large-scale Clinical Grade CAR T Cells

To prepare for in-house point-of-care (POC) CAR T manufacturing, a multidisciplinary ecosystem was established with support from multiple departments, as outlined in **Figure 2**. Large-scale CAR T production was performed using the CliniMACS Prodigy system with fresh leukapheresis and the TS520 tubing set (**Figure 3A**). A sample of the apheresis product was analyzed using the Sysmex automated hematology analyzer to determine leukocyte and lymphocyte counts, as well as red cell contamination, (**Figure 3B**) Sample was also sent for microbial testing and immunophenotyping. CAR T cells were manufactured using the TCT software program with all clinical-grade reagents. All processing was conducted at the ISO 7 CAR T cell laboratory of King Faisal Specialist Hospital & Research Center. To initiate the culture process, T cells were suspended in TexMACS medium supplemented with human AB serum and IL-2. TransACT reagent was added to stimulate the T cells in the Prodigy cell culture chamber. Key parameters assessed included expansion rate, transduction efficiency, viability, purity, and functionality. During the expansion phase, cell growth and morphology were monitored using an internal microscope camera. CAR T cells formed typical clusters that grew over time (day 6 to day 9, the day of cryopreservation) (**Figure 3C**). Transduction was performed using a CD19 second-generation CAR construct with a third-generation lentiviral vector backbone (provided by Lentigen/Miltenyi Biotech) **(Figure 4A**). The vector was introduced by sterile welding of the supply bag to the access tubing. In-process parameters were set for process optimization and release specifications (**Table 1**), based on previous experience (9, 10) and in-process testing as outlined in **Table 2**. The 8-day manufacturing process demonstrated that sufficient numbers of CAR T cells required for patient infusions were achieved, with significant expansion from day 1 to day 9 (**Figures 4B and 4C**). The drug substance was defined as autologous donor/patient-derived T-cells, some of which were transduced to express the CD19 CAR transgene, termed CD19 CAR T cells. The drug product (DP)/Advanced Therapy Investigational Medicinal Product (ATIMP) was defined as autologous ALL Lentigen CAR T cells in their excipients, in the final container closure system, ready for infusion (**Figure 2**). The DP excipients included DMSO, HAS, and CliniMACS PBS/EDTA buffer, as shown in **Table 3**. This combination of excipients was optimized (9, 13) for the successful cryopreservation of ALL Lentigen CAR T cells and has been previously administered to human subjects (6, 9). The-ALL Lentigen CAR T cell product is a cryopreserved cell suspension using a controlled-rate freezer. The cryopreserved ATIMP cells were stored in vapor phase liquid nitrogen (≤ −130°C) until use.

**Figure 4.**
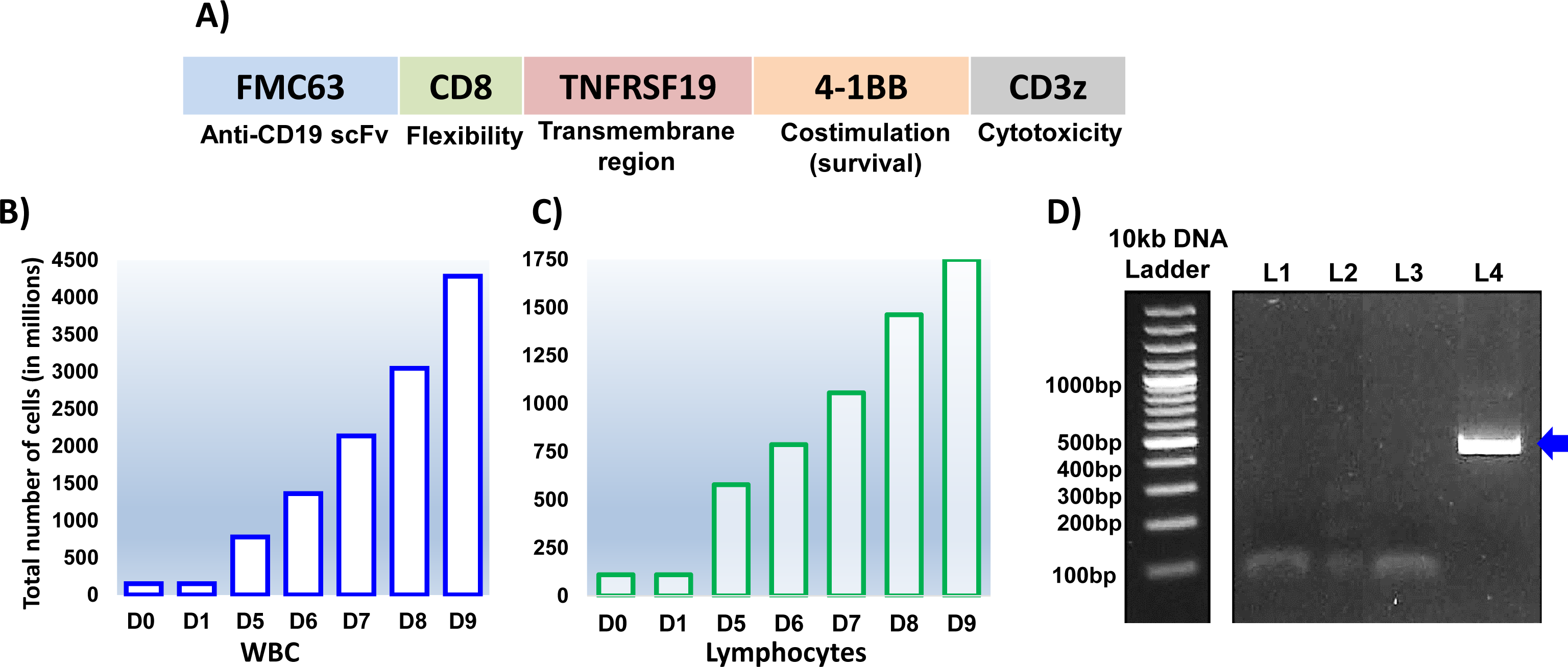
A) Schematic Representation of the Second Generation CD19 CAR Construct. The CAR sequence contains a single-chain fragment variable (scFv) derived from mouse anti-human CD19 antigen (clone FMC63), CD8 (hinge region), TNFRSF19 (tumor necrosis factor receptor superfamily, member 19), CD8-derived hinge region, TNFRSF19-derived transmembrane region, 4-1BB/CD137 costimulatory domain, and CD3-zeta chain intracellular signaling domain. **B) & C)** Cell expansion results show a 30-fold and 16-fold increase in total WBC and total lymphocytes count, respectively. **D)** Mycoplasma PCR test results for un-transduced T cells and CAR-transduced T cells. Lane 1 (L1): Un-transduced T cells; Lane 2 (L2): CAR T cells; Lane 3 (L3): Negative control; Lane 4 (L4): Positive control. The arrow indicates the expected target DNA band (500 bp) amplification in the positive control lane.

**Table 2.** In process parameters and controls for CD19 CAR T cells manufacturing process.

| Manufacturing Step | Day of Manufacture | Process Control/Specification |
| --- | --- | --- |
| Preparation of process media bag 1, bag 2 and buffers, TCT TS520 tubing installation and integrity check | d-1 or fresh cells | **Sterility of cells (BacT/ALERT + gram staining)\***<br>Cell count of starting material (CBC by Sysmex)<br>Automatic testing tubing-set integrity |
| Pre-CD4/CD8 selection on CliniMACS Prodigy<br>Preparation of leukapheresis | d0 for fresh cells | Pre-Selection **0.5 - 3 X10<sup>9</sup> WBC\***<br>**Sterility of cells (BacT/ALERT + gram staining)\***<br>Immunology (FACS)<br>Pre-CD4/CD8 selection cell count (FACS)<br>Viability (FACS) |
| Post-CD4/CD8 section on CliniMACS Prodigy<br>Preparation of Cells | d0 for fresh cells | Immunology (Basic Panel)<br>Post-CD4/CD8 selection cell count (FACS)<br>Viability (FACS)<br>Culture Set up Post-Selection **70-200 X10<sup>6</sup> \* CD3+ T Cells\*** |
| Culture setup preparation of CliniMACS Prodigy | d0 | Cell loading and activation of cells |
| Culture Set-Up/<br>Start of TCT | d0 | Activity matrix setup and gas mix setup |
| Transduction | d1<br>d6 | Activation of cells assessment (optional)<br>**Sterility of cells (BacT/ALERT)\***<br>**Activation marker expression on day 1 by measuring early (CD69), middle (CD25), and late (HLA-DR) T-lymphocyte activation markers (Optional)\***<br>Preliminary CD19-CAR staining (FACS)<br>**≥15%\* Transduction CD45+ CD3+ CAR+ T Cells\***<br>Cell number >D5 |
| Expansion | daily: d4-d8 /12 | Cell count (CBC by Sysmex)<br>**Cell number >D5 >D6 >D7 >D8 / >D9, >D10 >D11\*** |
| Harvest and cryopreservation | d8 or d12 | **Total cell number required to produce the IMP dose: CD45+CD3+ CAR+ T Cells ≥100x10<sup>6</sup> \***<br>Sterility of cells (BacT/ALERT + Gram staining, Mycoplasma, Endotoxin testing)<br>VCN, RCL, CD3/CD28 bead testing and potency testing.<br>Immunology (FACS)<br>Cell count (CBC by Sysmex)<br>Viability (FACS) |

*\*These are critical steps in the manufacturing process. If the specification are not met for these critical steps (including sterility should be NO growth, no organisms seen) the manufacture may be terminated if necessary.*

**Table 3.** Excipients used in ALL Lentigen CAR T cell product.

| Excipient Components | Supplier | Origin | Certification |
| --- | --- | --- | --- |
| **CliniMACS PBS/EDTA buffer** | Miltenyi Biotec | Chemical | CofA/CofC/Product Quality Certificate GMP, ISO 13485, ISO 15747, ISO 10993-1, CE marked. |
| **20% Human Albumin Solution (Zenalb 20)** | Bio products Laboratory | Biological | Licensed medicinal product (PL 08801/0007)<br>Or Appropriate alternative equivalent licensed product |
| **CryoSure-DMSO** | WAK-Chemie Medical, GMBH | Chemical | CofA, CofC, CE marked, ISO 13485, USP Grade |

## 3. Process Related Impurities and Cell Phenotype in the Final Drug Product

Process-related impurities in the product may include microbial contaminants and endotoxins. These potential impurities were evaluated in the final cell product using gram staining, bacterial cultures, endotoxin assay (**Table 1**), and mycoplasma detection by PCR (**Figure 4D**). Additionally, potential carryover of process-related materials could include interleukin (IL2), residual human AB serum, TransAct reagent, and free vector particles. Specific measurements of interleukin and residual human AB serum carryover were not undertaken, as transduced T-cells undergo multiple washing stages during expansion to the final harvest, which are likely to eliminate such impurities. Experiments conducted by the manufacturer (Miltenyi Biotech) confirm that the carryover of TransAct in an 8-12 day process with a culture wash on days 3-12 is negligible (**Table 4**). Furthremore, TransAct b e a d s a r e biodegradable, therefore, they do not require removal prior to drug formulation. Therefore, the overall potential risk associated with the use of *In-Vitro* stimulated and expanded T cells is considered extremely low concerning potential direct effects of the MACS GMP TransAct CD3/CD28. T cells stimulated with MACS GMP TransAct CD3/CD28 are incapable of activating adjacent T cells. Furthermore, negligible viable lentiviral vector particles are expected in the final product, as these are unstable with a half-life at 37°C of approximately 350 minutes (14). Similar to other *Ex-Vivo* gene therapy products, residual impurities from lentiviral production, including residual plasmid DNA and 293T cell residue, may be present but are quantified during the vector release process. Cellular composition was analyzed using flow cytometry for the presence of CD45+ lymphocytes, CD3+ T cells, CD14+ monocytes, CD19+ B cells, natural killer (NK) cells, and CD3+CD56+ NK T cells on day 0 and day 8 (day of cryopreservation). There was a clear difference in the phenotype of the transduced and expanded final drug product compared to the starting material cell composition. The active drug substance is autologous T-cells, some of which are transduced to express the CD19 CAR transgene. The composition of the final product in terms of other blood cells, such as NK T cells and NK cells, is recorded in **Table 5**. Other cells may remain in the drug substance but are not considered harmful in the autologous setting. CliniMACS buffer, human albumin solution (HAS), and DMSO constitute the excipient. CliniMACS buffer is commonly used with cell products processed in CliniMACS systems; HAS is pharmaceutical grade, and DMSO is routinely administered with cryopreserved cellular products such as hematopoietic stem cells and donor lymphocyte infusions. This combination of excipients has been optimized and used previously for allogeneic and autologous CAR T cells (6, 9, 10, 13).

**Table 4.** In-house CAR T process on CliniMACS Prodigy and TCT work flow. Upon starting the process, an “activity matrix” is programmed by the user by the addition of different activity modules into the matrix. This makes the TCT process adaptable depending on the cell source, vector-type and desired expansion period for the respective product. There are three types of shaking that can be implemented in the Centricult unit depending on the cell concentration in the chamber: Type 1, 100 rpm, one direction, recommended >2 × 10^6^ cells/mL; Type 2, 300 rpm, one direction, recommended >3 × 10^6^ cells/mL; Type 3, 300 rpm, change of direction, recommended >4 × 10^6^ cells/mL.

| Day | Activity Modules (Parameters) | Volume (in mL) |
| --- | --- | --- |
| **0** | Start of Culture | 70 |
| **1** | Transduction (reagent vol.: 10) | 100 |
| **3** | Culture wash (1 cycle) | 200 |
| **3** | Activate shaker (Type 2) | 200 |
| **5** | Feed (port: 3, vol. (+): 50) | 250 |
| **6** | Media exchange (port: 3, vol. (-/+): 125/125) | 250 |
| **6** | Activate shaker (Type 3) | 250 |
| **7/8** | Media exchange (port: 3, vol. (-/+): 150/150) | 250 |
| **8** | End of culture (optional) | 250 |
| **9 / 10 / 11** | Media exchange (port: 3, vol. (-/+): 180/180) | 250 |
| **12** | End of culture | 250 |

**Table 5.** Phenotype of different cell type composition in the final drug product.

| Cell Type | Day 0 (manufacturing start day) | Final product (before cryopreservation) |
| --- | --- | --- |
| **CD3+ in viable CD45+** | 58.38% | **98.84 %** |
| **NK T cells** | 6.30% | **5.85 %** |
| **NK cells** | 0.58 % | 0.72% |
| **Monocytes** | 19.74% | **0 %** |
| **Eosinophils** | 6.11% | **0.07 %** |
| **Neutrophils** | 5.72% | **0 %** |
| **B cells** | 9.21% | **0 %** |

## 4. Biological and Immunological Characteristics of CAR T cells

Clinical-grade CD19 CAR T cells, produced using the CliniMACS Prodigy system, were characterized in parallel with small-scale un-transduced control T cells for quality control assessment. The composition of T cells in donor leukapheresis samples was initially 37.2% CD4+ T cells and 52.9% CD8+ T cells on day 0, which increased to 40.7% CD4+ and 54.6% CD8+ T cells by day 9. T cell activation was confirmed by the high expression levels of CD25+ and CD69+ on both CD4+ and CD8+ T cells (**Figure 5 A and B**). Transduction efficiency of CD3+ T cells, as measured by flow cytometry on day 6 and day 9, was consistently 35% CD3+ CAR+ T cells (**Figure 5 C**). The stability of cryopreserved ALL Lentigen CAR T cells was assessed, showing a post-thaw viability of 94.3% after six months and 12 months, compared to 99% pre-cryopreservation (**Figure 5 D**), indicating that the product remains active after 12 months. To corroborate the flow cytometry results, quantitative assessment of the average CAR vector copy number per transduced cell revealed 1.93 copies/cell (**Figure 6 A**). The 3rd generation CD19 CAR vector used in this study is not FDA-approved; hence, to ensure the absence of replicating virus in the final drug product, an RCL assay was performed to detect VSVg DNA copies. The RCL assay confirmed the absence of replication-competent lentivirus in the final product (**Figure 6 B**). Notably, RCL has never been observed in vector preparations made using the 4-plasmid system (15, 16). Additionally, all sterility assessments conducted pre-cryopreservation showed no microbial growth, indicating that the final drug product is free of microbial contaminants (**Table 1**).

**Figure 5.**
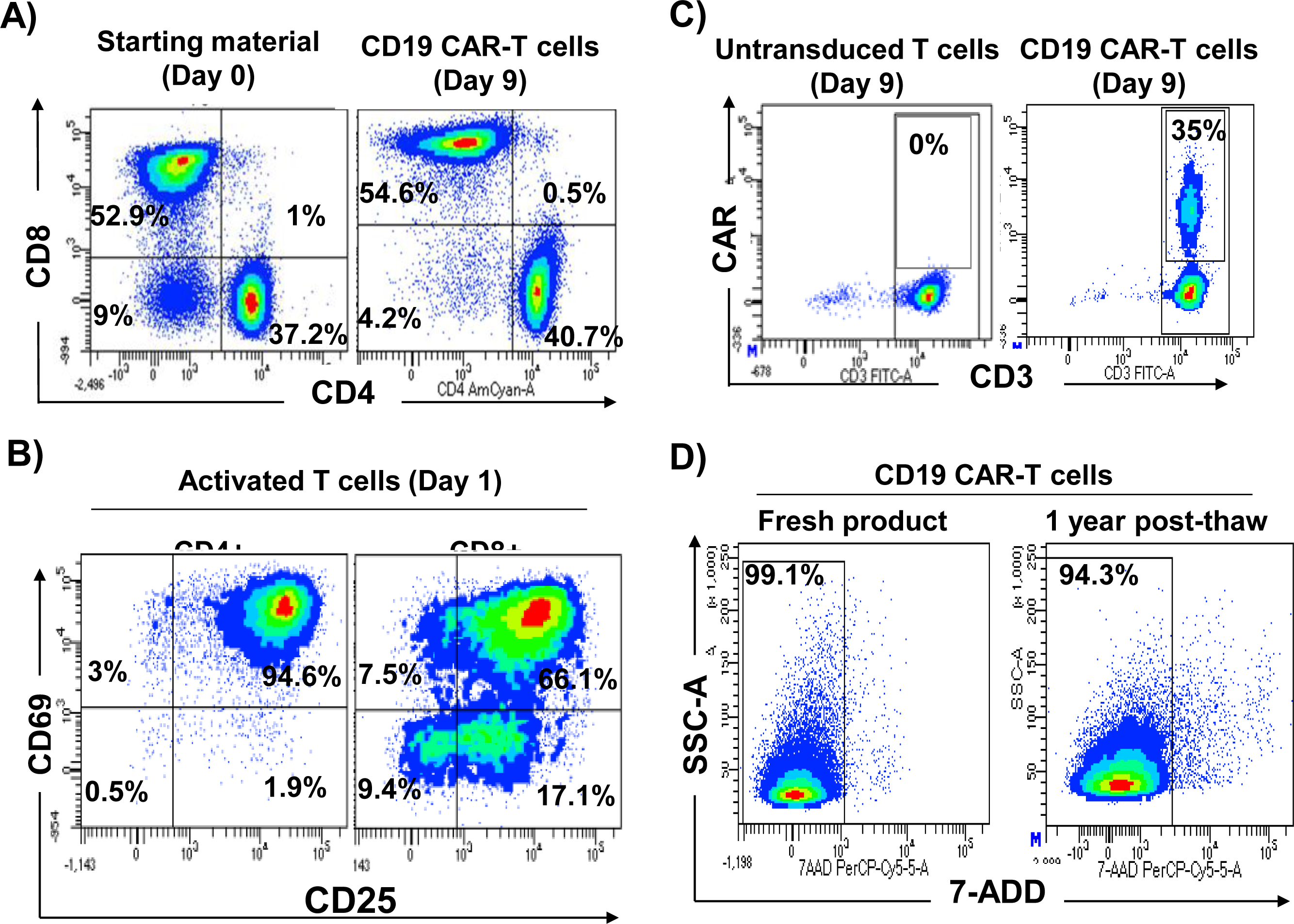
Clinical Grade CD19 CAR T cells Phenotyping and Assessment. **A)** Composition of CD3+ Cells. Flow cytometry was used to analyze the composition of CD3+ cells on day 0 and day 9 (end of culture). The cells were stained with antibodies against CD3, CD4, and CD8. **B)** T Cell Activation. 24 hrs post-activation with TransAct, T cells were stained with antibodies against CD3, CD4, CD8, CD25, and CD69. The percentages of CD25+ and CD69+ cells among CD4+ and CD8+ populations were measured using flow cytometry. **C)** CD19 CAR T Cells Transduction Efficiency. On day 9, the transduction efficiency of CD19 CAR T cells was determined by staining with CD3 and a CAR detection recombinant protein, followed by flow cytometry analysis. The percentage of CAR+ cells among CD3+ cells was recorded, with un-transduced T cells serving as a negative control. **D)** CD19 CAR T Cells Stability. The viability of CD19 CAR T cells was assessed using 7-AAD staining and flow cytometry on day 9 (pre-cryopreservation) and after one year of storage in liquid nitrogen (1 year post-cryopreservation).

**Figure 6.**
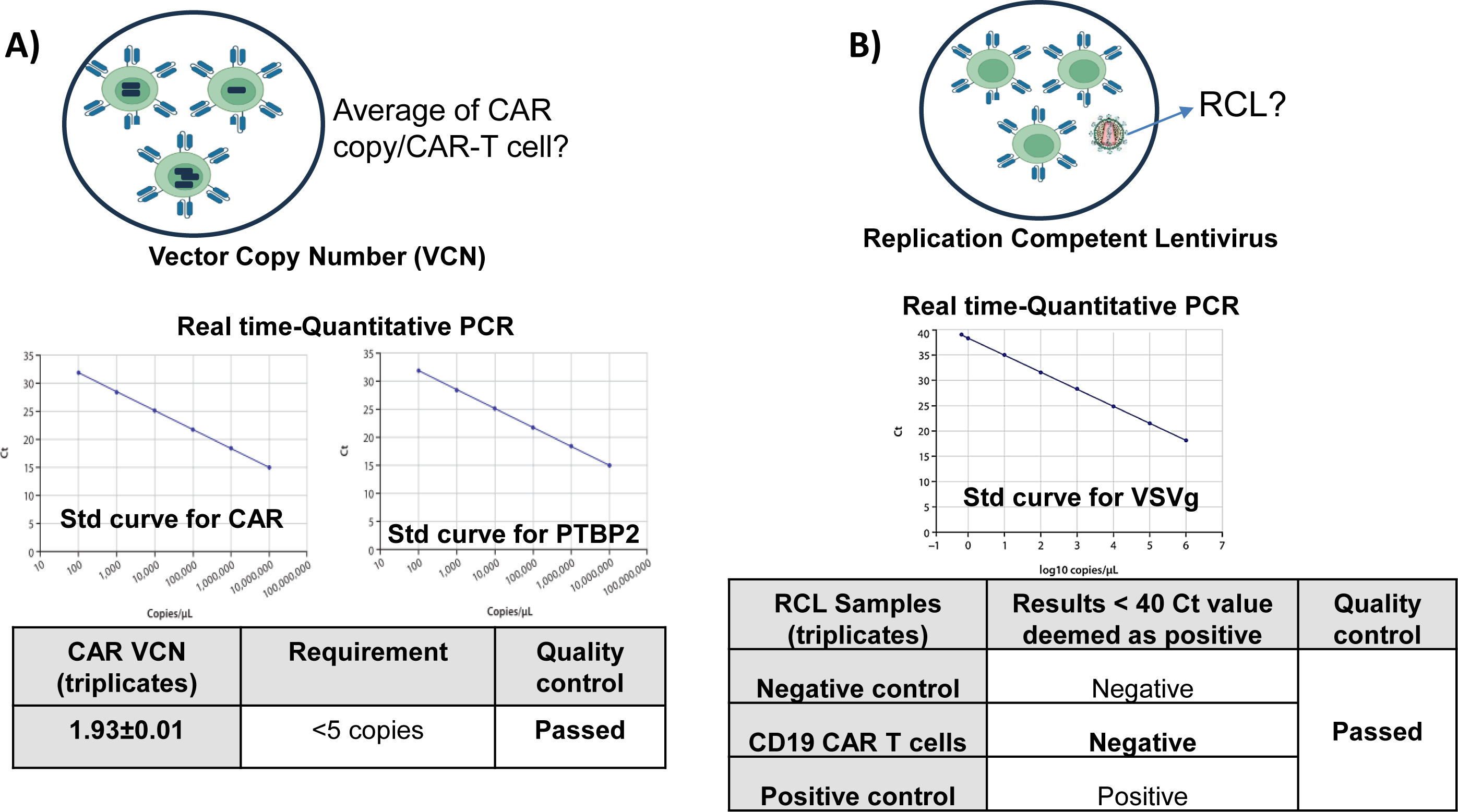
Real-Time Quantitative PCR. **A)** The Vector Copy Number (VCN) assay quantifies integrated lentiviral vector copies in transduced cell genomes. It involves extracting genomic DNA and performing real-time quantitative PCR (qPCR) with vector-specific primers. The VCN is calculated by comparing vector sequence amplification to a reference gene, providing an accurate measure of vector copies per cell. This is crucial for assessing gene transfer efficiency and safety, ensuring optimal therapeutic gene levels without risking insertional mutagenesis. **B)** The Replication Competent Lentivirus (RCL) Assay detects replication-competent lentivirus in gene therapy products, ensuring safety. It involves culturing transduced cells, collecting supernatants over time, and performing qPCR to identify any replication-competent sequences. Absence of amplification confirms the product is free from replication-competent lentivirus, ensuring its safety and integrity.

## 5. Clinical Grade CAR T cells Showed high Potency Against CD19+ Human Cell Lines

To evaluate whether the automated expanded clinical-grade CD19 CAR T cells retained their functionality against CD19-expressing target cells, post-thaw co-cultures were conducted using these T cells with CD19+ Raji and Daudi (B-cell-derived leukemic cell lines) as target cells, and K562 cells as a negative control (CD19-target cells) (**Figure 7 A**). As illustrated in **Figures 7 B and C**, the clinical-grade CD19 CAR T cells exhibited significant cytotoxic effects after 24 hours of co-culture at various effector-to-target (E:T) ratios with Raji and Daudi cell lines, in contrast to K562 cells. Consistent with these findings, co-culture of the clinical-grade CD19 CAR T cells with Raji and Daudi (CD19+ cell lines) resulted in the secretion of IFN-γ, whereas co-culture of un-transduced control T cells with the same targets did not produce detectable levels of IFN-γ (**Figure 7D**). Additionally, intracellular cytokine staining revealed specific production of IFN-γ and TNF-α by CD19 CAR T cells after co-culture with Raji and Daudi, compared to un-transduced T cells (**Figures 8A and B**). CD19 CAR T cells co-cultured with CD19+ target cells secreted pro-inflammatory cytokines interferon-gamma (IFN-γ) and tumor necrosis factor-alpha (TNF-α) in response to their target antigen, whereas no cytokine secretion was observed upon co-culture with the control K562 (CD19-) cells (**Figures 7 and 8**). Collectively, these data confirm the functionality and specificity of the automated manufacturing process for T cells.

**Figure 7.**
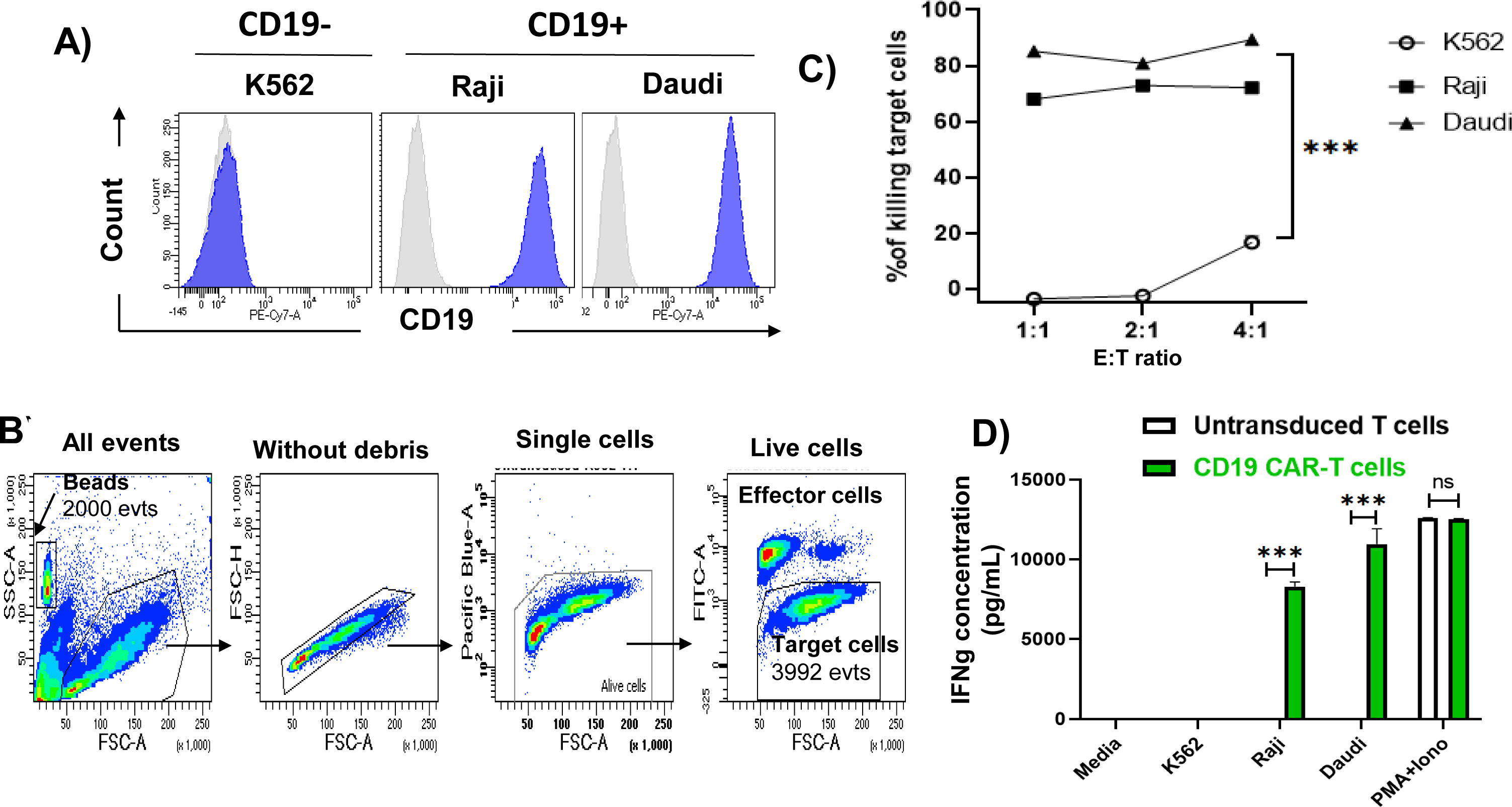
CAR T cells Demonstrate Effective Cytotoxicity Against CD19+ cells In-Vitro. **(A)** CD19 expression was assessed on the target cell lines Raji and Daudi using flow cytometry, with the K562 cell line serving as a CD19-control. **(B)** The gating strategy employed quantified the absolute number of live target cells utilizing absolute count beads (TruCount). **(C)** CAR T cells selectively and efficiently eradicated CD19+ cells at effector-to-target (E:T) ratios of 10:1 and 5:1, as determined by the absolute number of target cells compared to those cultured with un-transduced T cells. **D)** Co-culture of CD19 CAR T cells with Raji and Daudi (CD19+ cell lines) induced highly significant IFN-γ secretion with both the target cell lines. In contrast, co-culture of control effector cells (un-transduced T cells) with CD19+ targets resulted in undetectable levels of IFN-γ secretion. Statistical significance was indicated by p-values of less than 0.0001 (***), comparing un-transduced T cells to CD19 CAR T cells.

**Figure 8.**
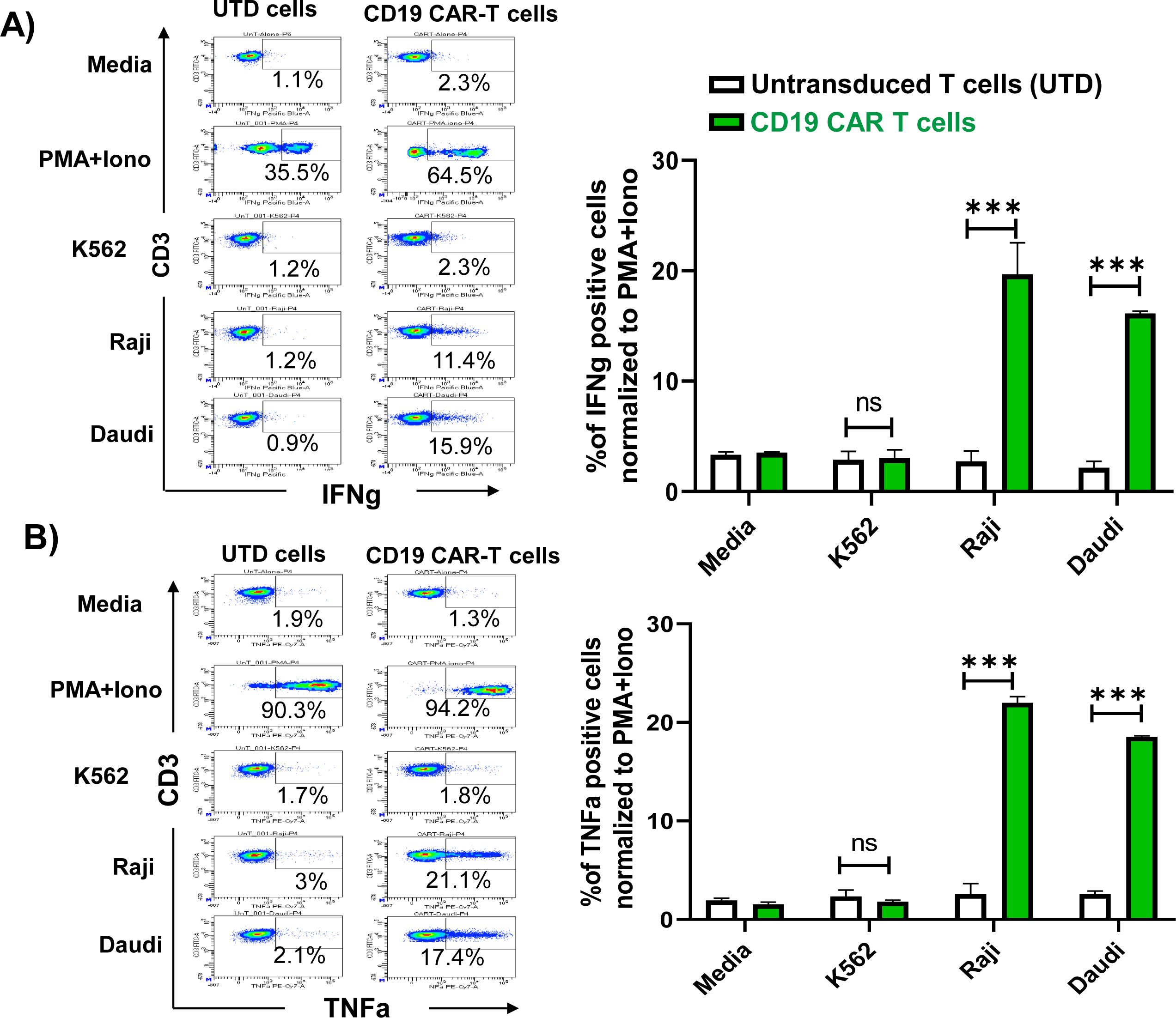
Intracellular staining for IFN-γ and TNF-α was conducted following the co-culture of CAR T cells and un-transduced T cells with CD19+ target cells (Raji and Daudi). After 6 hours of co-culture, cells were stained with antibodies against CD3, IFN-γ, and TNF-α. The K562 cell line was used as a CD19-target control. The percentage of IFN-γ+ **(A)** and TNF-α+ **(B)** cells among CD3+ cells was determined. Statistical significance was indicated by p-values of less than 0.0001 (***), comparing un-transduced T cells to CD19 CAR T cells.

## 6. Phenomenon of Trogocytosis in CD19 CAR T Cells

We observed an unexpected phenomenon of trogocytosis involving CD19 CAR T cells. Specifically, after co-culturing effector CAR T cells with CD19+ target cells (Raji and Daudi), we were unable to detect the CAR on the surface of CAR T cells using biotinylated recombinant CD19 protein. Notably, CAR expression remained unaffected when co-cultured with CD19-cells, as shown in **Figure 9A**. We hypothesize that this masking effect is due to the transfer of CD19 antigens from the CD19+ cell lines to the CAR T cells. To further explore this hypothesis, we proposed that CAR T cells might acquire membrane-bound CD19 proteins from CD19+ target cells during co-culture, as illustrated in **Figure 9B**. This acquisition could lead to the masking of the CAR T cells’ ability to bind recombinant CD19 protein. To verify our hypothesis, we stained the cells with DIO and DiL and conducted flow cytometry analysis. The results demonstrated a significant increase in trogocytosis-positive cells (DIO+DiI+) in direct contact with both, Raji and Daudi co-cultures compared to those co-cultured with K562 (CD19-), as shown in **Figure 9C**. This indicates a transfer of membrane components from CD19+ cells to the CAR T cells. Other did not observe trogocytosis phenomino using similar CD19 CAR vector (17–19). Therefore, understanding trogocytosis and its effects on CD19 CAR T cells is crucial for optimizing therapy and improving patient outcomes (20–22). This process can impact the function and efficacy of CAR T cells, and warrants further research into its molecular mechanisms.

**Figure 9.**
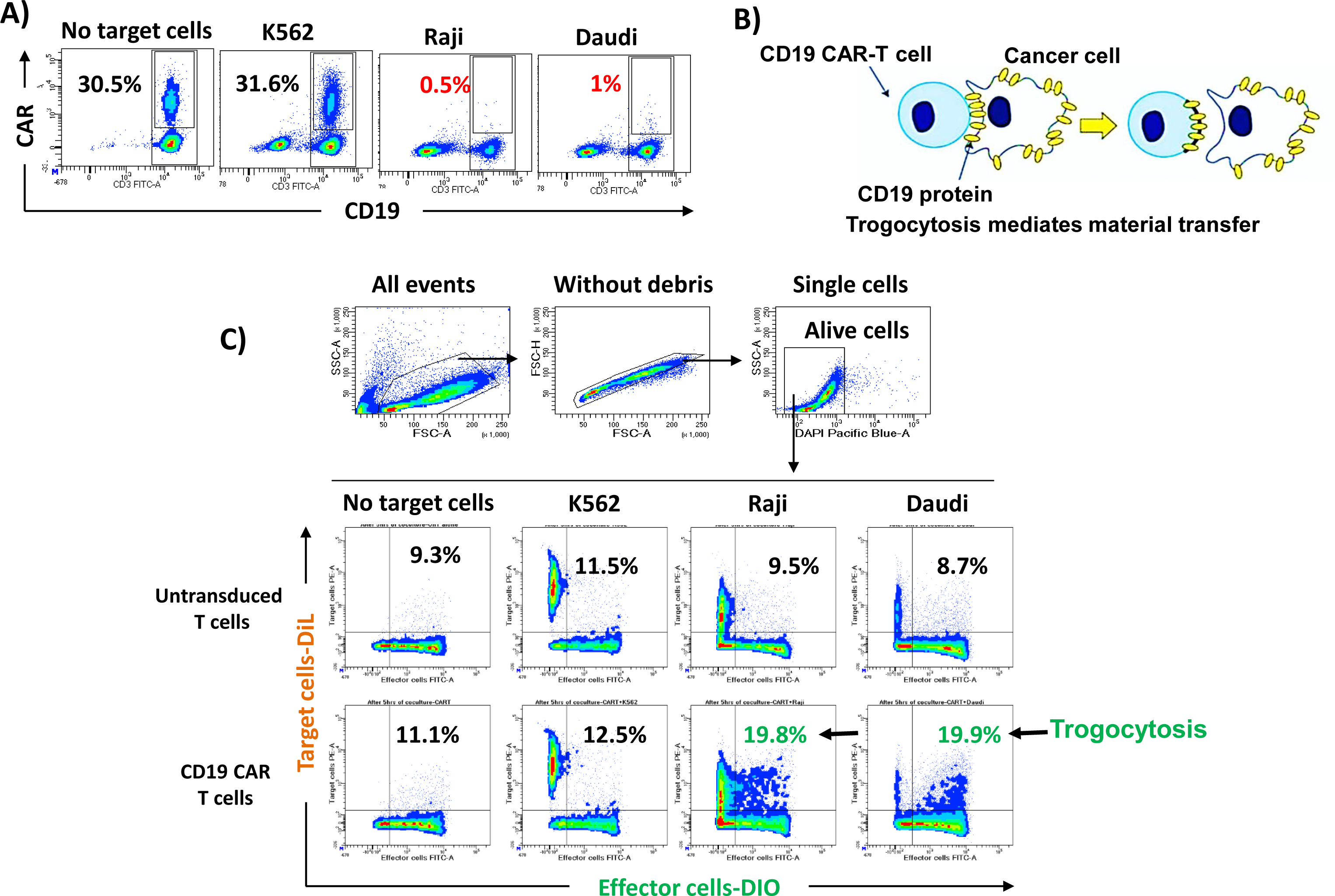
Evaluation of Trogocytosis Phenomenon. **A)** The expression of CAR on T cells (CD3+) was assessed using flow cytometry following co-culture of CAR T cells with target cells. The percentage of CAR+ cells among CD3+ cells is presented. **B)** Schematic illustrates the transfer of cell surface CD19 molecules, along with membrane patches, from donor cells to recipient cells (trogocytosis-mediated material transfer), adapted from Miyake and Karasuyama., 2021 (20). **C)** The gating strategy used to identify the percentage of trogocytosis-positive cells is described. Effector cells (both un-transduced T cells and CART cells) were labeled with DIO, while target cells (K562, Raji, and Daudi) were labeled with DiL prior to a 6-hour co-culture period.

## 7. Protein Expression Changes between Un-transduced T cells and CD19 CAR T cells sample groups

Whole cell lysates from Un-transduced T cells and CD19 CAR T cells sample were analyzed using liquid chromatography tandem mass spectrometry (LC-MS/MS). Over 1300 proteins were identified from all the sample groups of which 491 were significantly differentially expressed (≥ 2 to ∞ - fold change and p < 0.05) between Un-transduced T cells & CD19 CAR T. The 491 differentially expressed proteins between sample groups were subjected to unsupervised hierarchical cluster analysis with the heat map illustrating the expression changes of the 491 differentially expressed proteins (**Figure 10**). This detailed protein profiling enhances our understanding of the molecular differences between un-transduced T cells and CD19 CAR transduced T cells and guides further research into therapeutic mechanisms and target identification in CAR T cell therapy. The profound molecular distinctions induced by CAR expression in T cells could be pivotal for optimizing therapeutic strategies in cancer treatment. The data clearly highlights the profound molecular differences induced by CAR expression in T cell and could be pivotal for further investigations into the mechanisms of action and for optimizing therapeutic strategies in cancer treatment.

**Figure 10.**
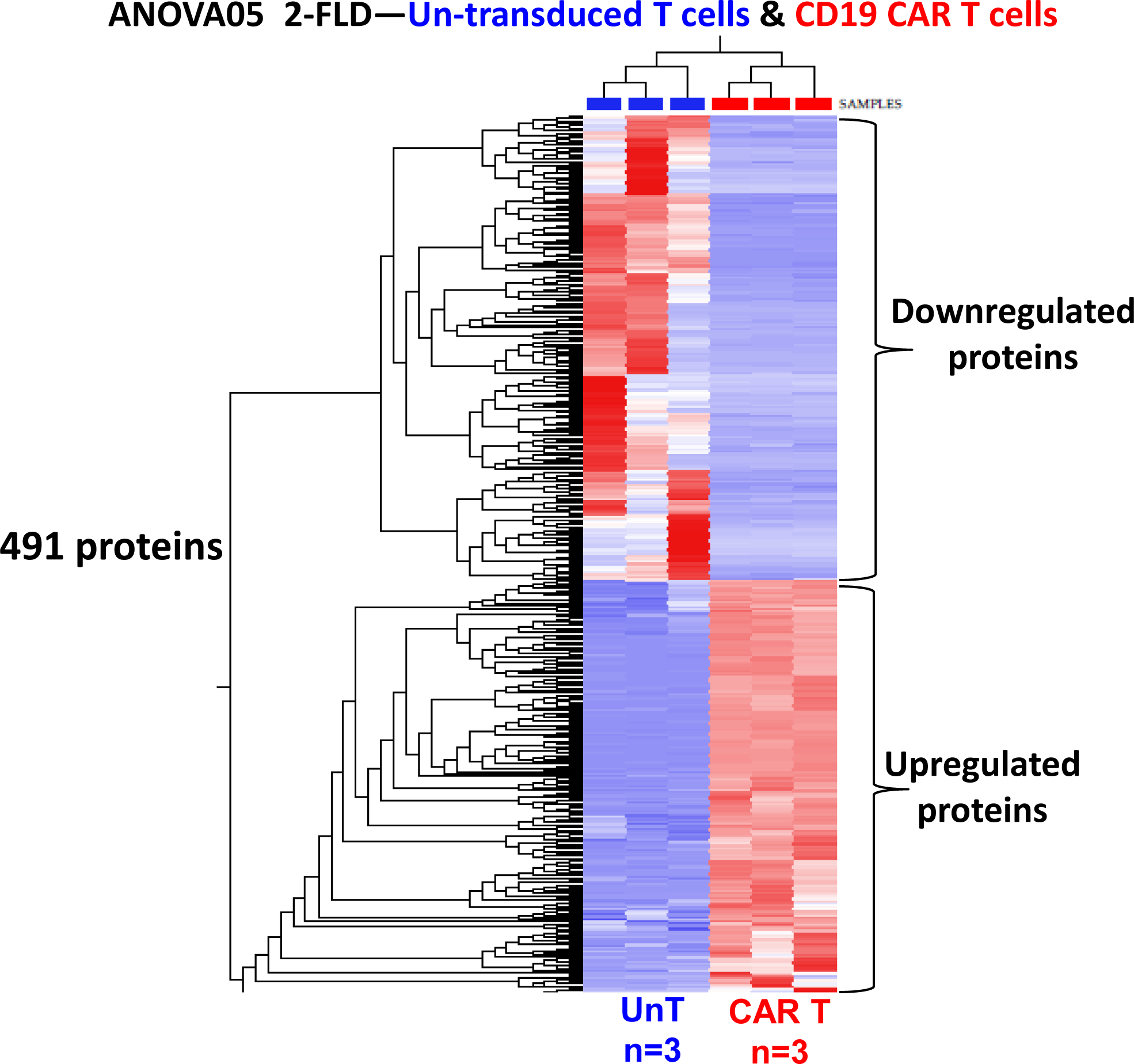
Differential Protein Expression Profile in Un-transduced and CD19 CAR Transduced T cells. Unsupervised hierarchical cluster analysis using the dataset of 491 differentially expressed proteins between Un-transduced T cells (UnT) & CD19 CAR T cells. The heat map show the relative measures of proteins as either upregulation in red color and downregulation in blue. The image was generated using the Qlucore Omics Explorer version 3.7 (Lund, Sweden) (https://qlucore.com). Details of these proteins including accession number, identified peptides, p values fold change and descriptions are as listed in **Supplementary Table S1**.

## DISCUSSION AND CONCLUSSION

Chimeric Antigen Receptor (CAR) T cell therapy has revolutionized cancer treatment, especially for patients with refractory or relapsed hematologic malignancies. Producing clinical-grade CAR T cells requires reproducible, scalable, and sterile processes, skilled personnel, clean-room facilities, infrastructure, and a robust quality management system. We have established the first point-of-care facility for large-scale, clinical-grade CAR T cell production in Saudi Arabia at KFSH&RC. This facility aims to enhance the accessibility, affordability, and efficiency of this therapy. Our central hypothesis is that local manufacturing of CAR T cells will significantly reduce production costs, alleviate logistical challenges, and expedite personalized treatments, thereby improving patient outcomes and fostering local biomedical expertise and infrastructure. This initiative positions Saudi Arabia as a regional leader in innovative cancer therapies and biomanufacturing. We demonstrated the feasibility of decentralized manufacturing of anti-CD19 CAR T cells using the Miltenyi CliniMACS Prodigy system. Manufacturing, conducted exclusively with CliniMACS Prodigy and Miltenyi Biotec reagents in the ISO7 air-handled CAR T Laboratory at KFSH&RC, was completed in 8 to 12 days. We successfully generated clinical-grade CAR T cells from healthy donor apheresis products, yielding 6 to 8 doses for a Phase 1 clinical trial without any production failures. These on-site produced CAR T cells can be used immediately (reducing vein-to-vein time) or cryopreserved for later use. The transduction efficiency was 35% with a viability of 98%, meeting all ATIMP release criteria and clinical application standards. The results showed a 30-fold expansion of CAR T cells. Cost analysis in this academic, not-for-profit setting estimates the cost per product at approximately 250K SAR (excluding workforce costs), potentially improving therapy access. Our findings underscore the safety and efficacy of this process, supporting its potential for future clinical trials.

In conclusion, we have established a GMP-compliant ISO7 facility for point-of-care CAR T cell production using the CliniMACS Prodigy device. This development could significantly enhance patient outcomes and advance CAR T cell therapy in Saudi Arabia. The facility oversees the entire manufacturing process, from leukapheresis to cell infusion, with bioreactors, cell processing units, and quality control systems designed specifically for CAR T cell production, ensuring high throughput and seamless integration. Standardized protocols and quality assurance measures were developed in line with international regulatory standards, in collaboration with the Saudi Food and Drug Authority (SFDA) to ensure compliance and facilitate approval. The production process was optimized to lower costs by sourcing raw materials strategically, using resources efficiently, and streamlining operations, thus reducing reliance on international suppliers. A comprehensive training program was initiated to build local expertise in CAR T cell manufacturing, covering technical skills, regulatory compliance, and quality control practices. Partnerships with stakeholders within KFSH&RC and international experts were established to develop a skilled workforce capable of sustaining and advancing the facility’s operations. A clinical application framework for CAR T cell therapy was created within the KFSH&RC healthcare system, including pathways for patient identification, enrollment, and follow-up. By integrating the manufacturing facility with academic and clinical research initiatives, we fostered a culture of research and innovation, supporting the continuous improvement of CAR T cell technologies and the development of next-generation therapies tailored to regional needs. These objectives aim to transform cancer treatment in Saudi Arabia, making CAR T cell therapy more accessible and positioning the Kingdom as a hub for biotechnological innovation and excellence. This effort aligns with Saudi Arabia’s Vision 2030, aiming to improve patient outcomes while contributing to economic diversification and healthcare advancement.

## AUTHOR CONTRIBUTIONS

R.E. F and F.S. are the principal investigator of this study. Simulation run design and experimental plan: F.S., W.W. Design of ISO7 lab, equipment procurement: F.S., R.E.F., N.A., S.O.A. Plan performed research: F.S., R.E.F., W.W., N.A., M.A., N. H., H.A., A.A., S.A., A.A., A.O.D. Full manuscript preparation: F.S., W.W. Review data and review paper: All the authors. Staff training for QA, QMS: F.S. Staff training for CAR T cell production: F.S. CAR T documentation and QC preparation: F.S., R.E.F, W.W., N.A., H.A, and S.O.A

## Data Availability

All data produced in the present work are contained in the manuscript

## ACKNOWLEDGMENTS

We gratefully acknowledge the generous and continuous support from 663 the higher management and the MCA for funding this project. Our sincere thanks extend to the various stakeholders at KFSHRC for their unwavering support, which has been crucial to the success of this project. We appreciate the collaborative efforts from multiple departments,including DPLM (Blood Bank, Microbiology, Toxicology, and stem cell laboratories) Center for Genomic Medicine (CGM), Environmental Monitoring, R&I, infection control& Hospital Epidemiology, HITA, Clinical Engineering, Supply Chain, and Purchase Departments, for their indispensable and ongoing contributions. Additionally, we express our gratitude to Lentigen, a Miltenyi Biotec Company (Gaithersburg, MD, United States), for kindly providing the CD19 CAR vector and reagents for this study.

